# Limits of lockdown: characterising essential contacts during strict physical distancing

**DOI:** 10.1101/2021.03.12.21253484

**Authors:** Amy Thomas, Leon Danon, Hannah Christensen, Kate Northstone, Daniel Smith, Emily Nixon, Adam Trickey, Gibran Hemani, Sarah Sauchelli, Adam Finn, Nicholas Timpson, Ellen Brooks-Pollock

## Abstract

COVID-19 has exposed health inequalities within countries and globally. The fundamental determining factor behind an individual’s risk of infection is the number of social contacts they make. In many countries, physical distancing measures have been implemented to control transmission of SARS-CoV-2, reducing social contacts to a minimum. Characterising unavoidable social contacts is key for understanding the inequalities behind differential risks and planning vaccination programmes. We utilised an existing English longitudinal birth cohort, which is broadly representative of the wider population (n=6807), to explore social contact patterns and behaviours when strict physical distancing measures were in place during the UK’s first lockdown in March-May 2020. Essential workers, specifically those in healthcare, had 4.5 times as many contacts as non-essential workers [incident rate ratio = 4.42 (CI95%: 3.88–5.04)], whilst essential workers in other sectors, mainly teaching and the police force had three times as many contacts [IRR = 2.84 (2.58–3.13)]. The number of individuals in a household, which is conflated by number of children, increases essential social contacts by 40%. Self-isolation effectively reduces numbers of contacts outside of the home, but not entirely. Together, these findings will aid the interpretation of epidemiological data and impact the design of effective SARS-CoV-2 control strategies, such as vaccination, testing and contact tracing.

## MAIN

The novel coronavirus, severe acute respiratory syndrome coronavirus 2 (SARS-CoV-2) causes a severe respiratory disease, termed COVID-19 [1]. The virus, first reported in the Hubei province of China in December 2019, spread rapidly across the globe [2]. SARS-CoV-2 is an airborne infectious disease transmitted between persons in close contact through respiratory droplets [3] [4]. Physical distancing measures have been implemented in many countries to reduce transmission, often at substantial economic and social cost. In the short-term, this has been a successful mitigation strategy, but vaccination and effective drug treatments are needed for long-term sustainable prevention of transmission and disease.

Quantifying the way that people interact, and the networks they form is important for understanding the speed and extent of infectious disease spread [5] [6]. Mathematical models that predict the impact of proposed interventions, often rely on data from social contact surveys to inform realistic transmission parameter choices [7] [8] [9]. In turn, determining the effectiveness of many interventions also relies on accurate and timely contact pattern information [7]. Moreover, rich participant data can help identify those who may be at increased risk of infection or identify those more likely to be involved in transmission events.

To reduce the number of social contacts, the UK government implemented strict physical distancing measures on March 23^rd^. Individuals were instructed to stay at home and only leave for essential work, daily exercise or purchasing of essential food/medicines. Many establishments were closed, including schools (open only to children of essential workers or those deemed vulnerable), non-essential shops, bars, restaurants, and sporting and entertainment venues. Only one published UK survey captured social contact patterns during the first COVID-19 lockdown [7]. Consequently, we have limited understanding on how social demographics and behaviours may influence contact patterns during times of physical distancing.

To address this data gap, we rapidly deployed a COVID-19 questionnaire to participants enrolled in the Avon Longitudinal Study of Parents and Children (ALSPAC): a prospective population-based cohort study in Avon, UK which recruited pregnant women in 1990–1992. This unique three-generational study comprises of three cohorts who have been followed for the last 30 years - a wealth of biological, genetic and phenotypic data has been collected [10] [11] [12] [13]. Utilising this resource, we aimed to quantify and investigate social contact patterns in the ALSPAC cohorts during the COVID-19 epidemic, when the first physical distancing restrictions were in place.

## METHODS

### Ethics

Ethical approval for the study was obtained from the ALSPAC Ethics and Law Committee and the Local Research Ethics Committees. Informed consent for the use of data collected via questionnaires and clinics was obtained from participants following the recommendations of the ALSPAC Ethics and Law Committee at the time. Participation was voluntary and analyses were performed on anonymised data.

### Cohorts

ALSPAC is an intergenerational prospective birth cohort from the southwest of England. The study recruited 14,541 pregnant women with expected dates of delivery between 1^st^ April 1991 to 31^st^ December 1992 in the county of Avon (eligible sample). This cohort of original pregnant women, the biological fathers and other carers/partners are known as the ‘G0’ cohort. Of the pregnancies initially enrolled, 13,988 children were alive at 1 year of age. When the oldest children were approximately 7 years of age, this initial sample was bolstered with cases who would have been eligible to join the study but originally failed to. Following this additional recruitment, 14,901 children were alive at 1 year of age [10] [11] [12]. This cohort of index children are known as the ‘G1’ cohort and have been followed since birth with measures obtained through clinical visits and questionnaires. The G1 cohort now includes their partners, referred to as G1 partners of offspring; the cohort of offspring of these index children are known as ‘G2’ [13]. Full details of the cohort and study design have been described previously and are available at http://www.alspac.bris.ac.uk.

### Data collection

An online questionnaire was developed to capture information on COVID-19 infection and behaviours in ALSPAC participants when physical distancing measures were first implemented in the UK, known as ‘lockdown’. The questionnaire was launched on 9^th^ April 2020, shortly after the announcement of official lockdown in the UK on 23^rd^ March. All participants enrolled in the ALSPAC G0 and G1 cohorts for whom we had a valid email address were invited to complete the questionnaire. Invitees were emailed a reminder two weeks after the original invite went out. The questionnaire was deployed and hosted using REDCap (Research Electronic Data CAPture tools) [14] [15], running for 5 weeks until 15^th^ May 2020. Full details on the questionnaire development and deployment can be found in the Wellcome Open Research data note [16].

The questionnaire was comprised of four sections recording information on: a) general health, recent travel, COVID-19 and/or influenza-like-illness symptoms; b) self-isolation, activities and contacts; c) pandemic impact (worries and feelings); d) accommodation and household structure, keyworker and healthcare worker (HCW) status, COVID-19 awareness/knowledge. The contact section of the questionnaire was based on that used in [17]. Participants were asked one question: “How many people, apart from those you live with, did you speak to yesterday in the following ways (for personal and for work reasons)?” Participants were asked to record four types of contacts: face-to-face (in person); over the phone; via video media; and physical (skin-to-skin touching). Participants recorded the number of each contact types in four different age groups: 0–4; 5–17; 18–69; and ≥ 70 years of age. Participants’ sex, date of birth and ethnicity (defined as ‘White’ or ‘Non-white’) were collected at the time of enrolment into ALSPAC. Information on participant occupation was collected from a separate questionnaire conducted in December 2019 as part of the ALSPAC regular data collection exercise. Occupational titles were coded according to the Standard Occupational Classification 2010 Coding Index [18]. Current address was assumed to be that recorded in the ALSPAC administrative database as of April 2020. Please note that the study website contains details of all the data that is available through a fully searchable data dictionary and variable search tool (http://www.bristol.ac.uk/alspac/researchers/our-data/).

### Data analysis

The number of daily contacts, consisting either of face-to-face or physical contacts per person was measured. Contacts involving physical touch were considered independent of those with face-to-face contact only. The total number of daily contacts per person was taken as the sum of face-to-face and physical contacts. The average number of contacts (face-to-face, physical and total) were stratified by age, sex, cohort, household size, presence of children in household (household composition), day of the week, occupation and self-isolation status. For age, participants were categorised into six age bands: 23–29; 30–39; 40–49; 50–59; 60–69; and 70+. We analysed answers related to symptoms and behaviours descriptively. We present the number and percentage or mean and standard deviation where appropriate.

We investigated the occupational profile of questionnaire participants self-identifying as essential and non-essential workers. Participants self-identified according to government guidelines [19] as either: a keyworker; a healthcare worker (HCW); or neither (classified as ‘Other’). We re-classified participants identifying as both keyworkers and HCWs as HCWs only to give mutually exclusive groups used in subsequent analyses. Next, we mined available occupational data categorised according to ONS occupational groups (available through data linkage described above) to determine the top five group job titles for participants identified as HCWs, keyworkers and other (non-essential) workers. The frequency of reported COVID-19 symptoms and those of influenza like illness were calculated.

We assessed representativeness of the study sample by comparing the distributions of variables (sex, age, ethnicity) observed for participants to those in south west England and the UK using the UK Office for National Statistics (ONS) estimates (2019 ONS mid-year estimates [20]; 2011 census data [21]).

We examined the association between covariates (age, sex, household size, presence of children in household, occupation, day of the week and self-isolation status) and numbers of daily total contacts, electing to use a negative binomial regression appropriate for the observed data (Supplementary Figure 2). Motivated by the high proportion of zero counts (41.5%; 2825/6807) observed, we contrasted the negative binomial regression analysis with its zero-inflated version. A lower AIC score was observed for the zero-inflated (29124) model vs the negative binomial (29263). When the predicted number of zero counts were inspected, both models performed similarly and when plotting the observed vs predicted counts, the models almost overlapped with each other (Supplementary Figure 2). The more parsimonious negative binomial model was used in further analyses. R version 4.0 was used for all analyses.

## RESULTS

### Responses and participants

Invitations to participate were sent to 12,520 individuals. The questionnaire was completed by 6807 (54%) participants between 9th April 2020 and 14th May 2020. The bulk of responses occurred in two waves, each one week in duration: the first 7 days (3981/6807; 58.5%) and the second 10 days later (1487/6807; 21.8%). Peaks in responses coincided with the invitation and reminder emails (Supplementary Figure 1). Most contacts were reported for interactions occurring on weekdays (5016/6807; 73.7%) versus the weekend (1791/6807; 26.3%).

Participants were aged between 23 and 81 years of age and belonged to four ALSPAC participant types: G0 mothers, G0 fathers/partners, G1 offspring and G1 partners of offspring. Participants were mostly female (4895/6807; 72%) and either aged 23–29 (3039/6807; 44.7%) or 50–69 (3562/6807; 52.3%), reflective of the ALSPAC G1 and G0 cohorts respectively (Figure 1a). Most participants were white (6040/6807; 88.7%), with only 2% non-white (137/6807). Ethnicity was unknown for 9.4% of participants (630/6807). Participant characteristics (sex and age) were dissimilar to that of southwest England and the UK. Ethnicity was broadly similar to southwest England but dissimilar to the UK (Supplementary Table 1). We assumed 70% of participants lived in the Bristol/Avon area based on addresses recorded in the ALSPAC administrative database (as of April 2020 based on the G1 cohort); 3.7% (254/6807) of participants reported living outside the UK at the time of questionnaire completion.

**Figure 1:**
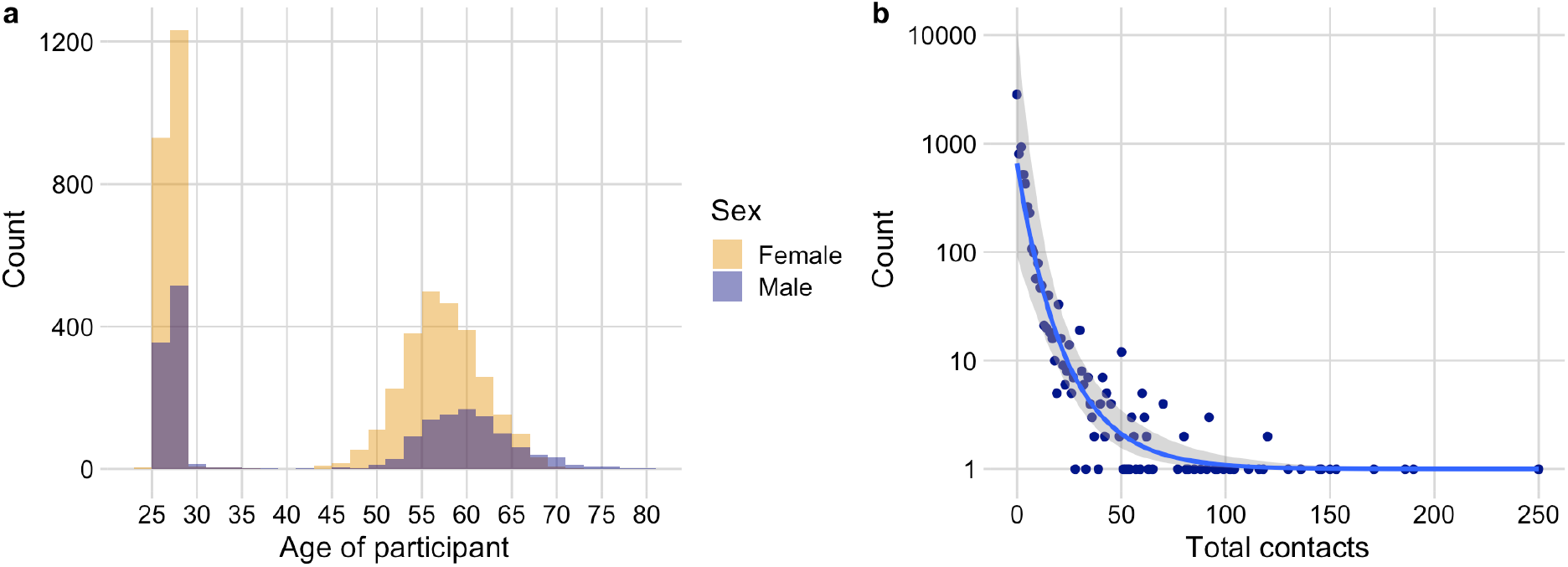
Distribution of participant age, sex and total contacts. a) Distribution of participant age coloured by sex. b) Frequency of total daily non-household contacts fitted to a negative binomial distribution (*NB(r=0*.*11, p=0*.*97)*) with 95% confidence interval shown.

### Associations with essential contacts

All participants recorded information on their daily contacts occurring outside of the household. At least one face-to-face or physical contact was recorded for 3944 (57.9%) and 900 (13.2%) participants respectively. No contact of either type was reported for 2825 participants (41.5%). Overall, a daily average of 3.7 [standard deviation (SD) = 10.6] total contacts outside the household were reported. The maximum number of face-to-face and physical contacts reported was 250 and 100 respectively. These contacts were reported for individuals aged between 23–29 years. Total contacts were higher on weekdays (mean = 4.0, SD = 11.4) compared to weekends (mean = 3.0, SD = 7.7).

Contacts were mostly face-to-face (mean = 3.4, SD = 10.0), with many fewer involving physical touch (mean = 0.3, SD = 2.1). We observed small differences in the number of contacts reported for G0 and G1 cohorts: on average, G0s reported increased face-to-face, but fewer physical contacts compared to G1s (Table ***1***). This observation was reflective of participant age, with a greater number of daily face-to-face contacts observed for individuals aged 50–59 (mean = 3.9, SD = 10.1) compared to participants aged 23–29 (mean = 3.4, SD = 11.3). Participants aged ≥ 70 years reported the fewest daily contacts (mean = 2.6, SD = 3.0).

**Table 1:** Daily contact numbers reported by participants (n=6807) stratified by age, sex, ALSPAC cohort, household size, household composition, day of the week, occupation and self-isolation status. For each age group, the mean (standard deviation) and number of participants are shown.

| Category | Value | Mean (SD) face-to-face contacts |  | Mean (SD) physical contacts |  | Mean (SD) total contacts |  | n |
| --- | --- | --- | --- | --- | --- | --- | --- | --- |
| Age (years) | 23–29 | 3.4 | (11.3) | 0.4 | (2.6) | 3.7 | (12.1) | 3039 |
|  | 30–39 | 4.3 | (15.6) | 0.8 | (3.8) | 5.1 | (17.3) | 45 |
|  | 40–49 | 5.9 | (16.7) | 0.7 | (2.7) | 6.6 | (17.7) | 90 |
|  | 50–59 | 3.9 | (10.1) | 0.3 | (1.6) | 4.2 | (10.5) | 2107 |
|  | 60–69 | 2.7 | (5.2) | 0.2 | (0.9) | 2.9 | (5.5) | 1455 |
|  | 70+ | 2.5 | (2.8) | 0.1 | (0.4) | 2.6 | (3.0) | 71 |
| Sex | Female | 3.5 | (10.1) | 0.3 | (1.9) | 3.8 | (10.7) | 4895 |
|  | Male | 3.2 | (9.6) | 0.3 | (2.5) | 3.5 | (10.3) | 1912 |
| Cohort | G0 | 3.5 | (8.7) | 0.2 | (1.4) | 3.7 | (9.1) | 3720 |
|  | G1 | 3.4 | (11.3) | 0.4 | (2.6) | 3.8 | (12.2) | 3087 |
| Household size | 1 | 2.7 | (7.7) | 0.1 | (0.9) | 2.8 | (7.9) | 712 |
|  | 2 | 3.4 | (10.0) | 0.3 | (1.4) | 3.7 | (10.4) | 3206 |
|  | 3 | 3.6 | (10.5) | 0.3 | (1.4) | 3.8 | (10.9) | 1554 |
|  | 4 | 3.8 | (11.7) | 0.6 | (4.5) | 4.4 | (13.8) | 859 |
|  | 5+ | 3.6 | (6.9) | 0.4 | (1.4) | 3.9 | (7.6) | 477 |
| Lives with children | Yes | 3.8 | (12.8) | 0.6 | (3.9) | 4.3 | (14.3) | 893 |
|  | No | 3.4 | (9.5) | 0.3 | (1.6) | 3.6 | (9.92) | 5914 |
| Day | Weekday | 3.7 | (10.9) | 0.3 | (2.1) | 4.0 | (11.4) | 5016 |
|  | Weekend | 2.7 | (6.8) | 0.3 | (2.0) | 3.0 | (7.7) | 1791 |
| Occupation | Health care worker | 8.2 | (14.3) | 0.9 | (4.1) | 9.1 | (16.4) | 697 |
|  | Keyworker | 5.8 | (16.5) | 0.4 | (2.8) | 6.1 | (17.0) | 1603 |
|  | Other | 1.8 | (3.8) | 0.2 | (1.0) | 2.0 | (4.0) | 4507 |
| Self-isolation | No | 3.8 | (10.8) | 0.3 | (2.1) | 4.1 | (11.4) | 5125 |
|  | Previously | 3.7 | (10.2) | 0.3 | (2.5) | 4.0 | (11.2) | 689 |
|  | Currently | 1.5 | (2.7) | 0.2 | (0.7) | 1.7 | (2.9) | 755 |
|  | Prefer not to say | 1.6 | (2.2) | 0.4 | (1.0) | 1.9 | (2.7) | 43 |
|  | Unknown | 0.3 | (2.3) | 0.005 | (0.072) | 0.3 | (2.4) | 195 |

### Household structures

The average household size was 2.6 persons (SD = 1.3, max = 29), with 10.6% (712/6807) of participants living alone. On average, household size was highest for the 40–49 age group (mean = 2.43, SD = 2.0, max = 11), thereafter household size decreased as participants aged (Figure 2a). The greatest heterogeneity in the total number of daily contacts was observed for households ranging from 1 to 4 persons in size (Figure 2c) – contact numbers tended to increase as household size increased. We observed increased household sizes to coincide with cohabiting with children aged 0 to 17 years old (Figure 2b). Adults living with children, on average, had 16% higher total contacts (Table ***1***) compared to those not living with children. Of note, those living with children had twice the number of contacts involving touch (mean = 0.6, SD = 3.9) compared to those living without children (mean = 0.3, SD = 1.6). Of the participant ages investigated, daily contacts were higher for participants aged 30–39 who lived with children compared to those who did not (Figure 2d).

**Figure 2:**
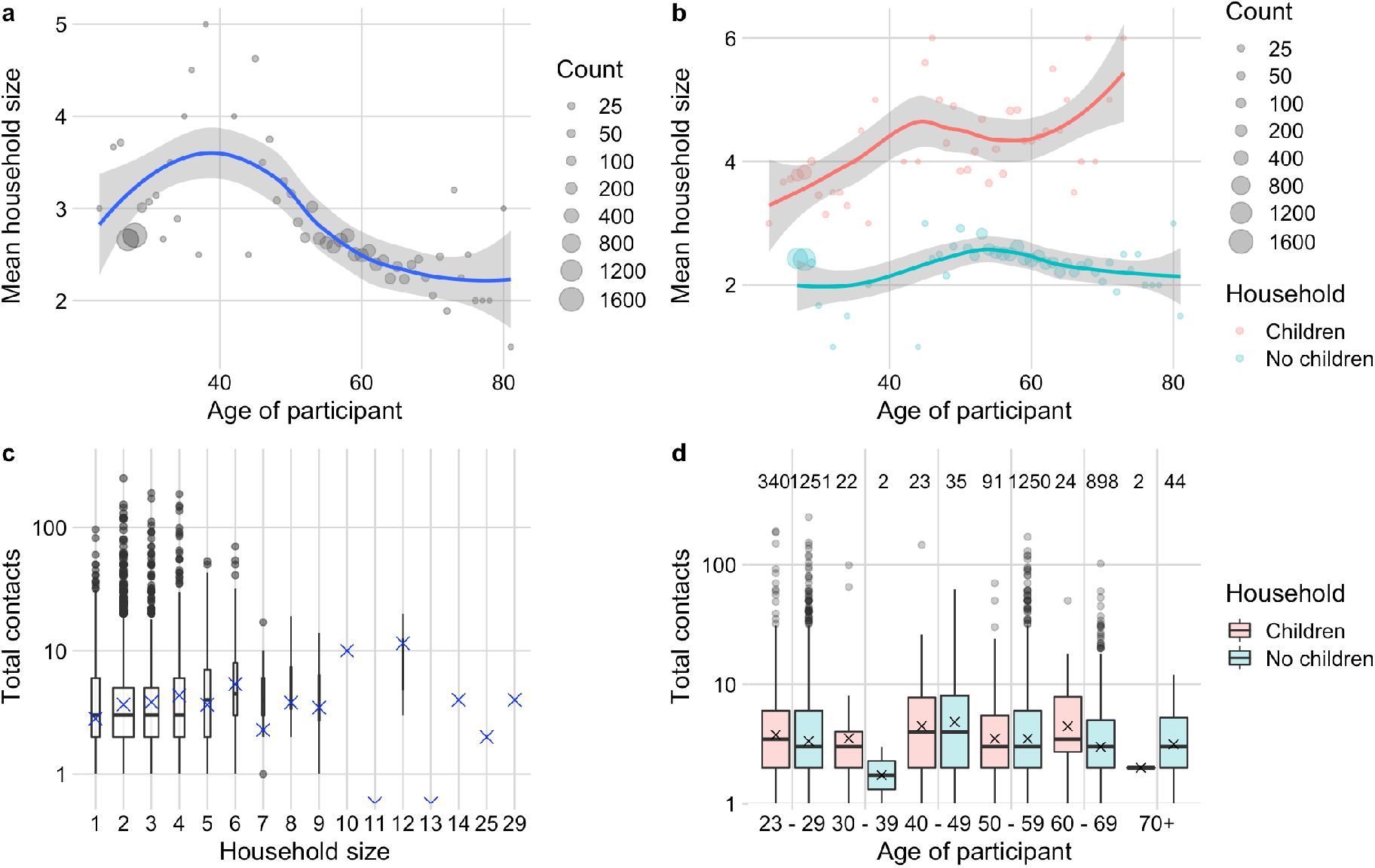
Participant household size and composition by number of contacts. a) Relationship between age of participant and mean household size. Grey points represent the mean household size at each participant age, with size proportional to the number of individuals contributing to the mean. The smoothed conditional mean with 95% confidence intervals is shown in blue. b) Effect of children in the household on mean household. Grey points represent the mean household size at each participants age, stratified by children in the household. Smoothed conditional mean with 95% confidence interval. c) Box and whiskers plot of the total daily contacts by household size; the middle line (hinge) corresponds to the median, the lower and upper hinges correspond to the interquartile range, and the whiskers extend to 1.5 times the interquartile range. The width of the boxplot is proportional to the number of observations. Outliers are shown as grey points and the geometric mean as red crosses. d) Box and whiskers plot of the total daily non-household contacts by children in the household (household composition). The number of observations contributing to each box is shown above.

### Occupation and health behaviours

We observed a clear occupational contact profile for HCW and keyworkers. Specifically, the number of face-to-face contacts and physical contacts appear positively correlated for HCWs. The majority of keyworkers had high numbers of face-to-face contacts but rarely reported physical contacts (Figure 3). There are outliers to this relationship: participants reporting a high number of face-to-face contacts only and individuals reporting a high number of physical contacts only. Mining available occupational group job titles showed nurses and medical practitioners were the largest groups of HCWs surveyed. Keyworkers were predominately teachers and school staff, with police officers identified as the fifth top group. The majority of participants not self-identifying as a HCW or keyworker (‘Other’) worked in office-based roles: administration, programming, sales, marketing and business (Table 2).

**Table 2:** Top five ONS group job titles for healthcare workers, keyworkers and other participants.

| **Healthcare workers** | **(n)** | **Keyworkers** | **(n)** | **Other** | **(n)** |
| --- | --- | --- | --- | --- | --- |
| Nurses | (67) | Primary and nursery teachers | (89) | Administrative work | (73) |
| Medical practitioners | (46) | Secondary education teachers | (27) | Programmers and software developers | (50) |
| Care workers and home carers | (21) | Nursery school staff | (26) | Sales | (46) |
| Physiotherapists | (12) | Teaching assistants | (22) | Marketing | (40) |
| Nursing assistants | (11) | Police officers | (21) | Business/Financial project management | (37) |

**Figure 3:**
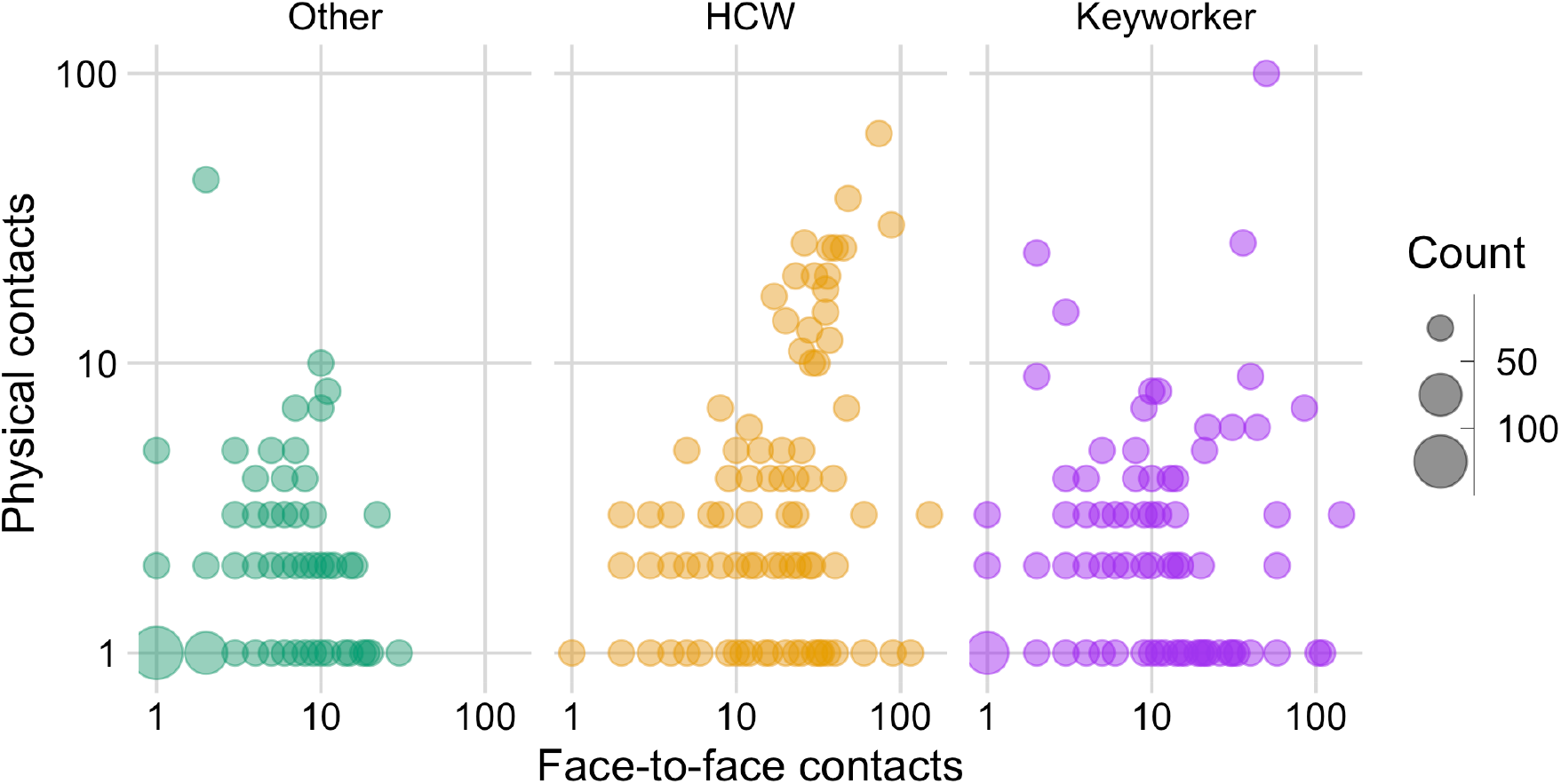
Total number of face-to-face contacts and physical contacts by occupation. Contacts stratified and coloured by occupational group: other (non-essential worker), healthcare worker (HCW) and keyworker. Size of the point is proportional to the count of individuals.

At the time of questionnaire completion, 11.1% of participants (755/6807) reported to be self-isolating, with a further 10.1% reported to have self-isolated previously (689/6807). The duration of self-isolation ranged from less than a week, up to more than 13 weeks. The majority of participants reported not to be currently self-isolating (5125/6807; 75.3%). We observed a 59% reduction in the average daily number of contacts for individuals self-isolating compared to those not (from 4.1 to 1.7) (Table 1). Some individuals preferred to not disclose their status (43/6807; 0.6%) or left the question blank (195/6807; 2.9%). Individuals who preferred to not disclose their isolation status had similar contacts to those currently self-isolating. Individuals who reported to self-isolate previously had similar numbers of contacts to those who had not self-isolated (4.0 vs 4.1).

Eighty-four participants (1.2%) had a confirmed positive test or were suspected to have had COVID-19 by their doctor. Of these cases, 8 were confirmed by a positive test result [16]. The top five symptoms most frequently reported were difficulty sleeping (20.1%), sneezing (18.3%), tiredness (14.7%), runny nose (12.8%) and sore eyes (10.2).

### Regression analyses of total number of contacts

Analysis of the total number of daily contacts with a multivariable negative binomial regression model shows a striking pattern of contact frequency by occupation – peak contacts were observed for healthcare workers, followed by keyworkers, with comparatively fewer contacts among non-essential workers (Table 3). Weekdays were associated with increased contacts compared to weekends. In the univariable model (Table 1), there was a rise in the number of contacts with age, peaking at 40–49 years and declining to the lowest number of contacts reported for adults aged ≥ 70 years. When controlling for covariates in the multivariable model, peak contacts occurred in adults aged over ≥ 70 years. Contacts were increased for females in the univariable model. Higher contacts were associated with living in a larger household and living with children. Self-isolation was associated with reduced contacts and for those that preferred not to disclose their isolation status or if the status was unknown.

**Table 3:**
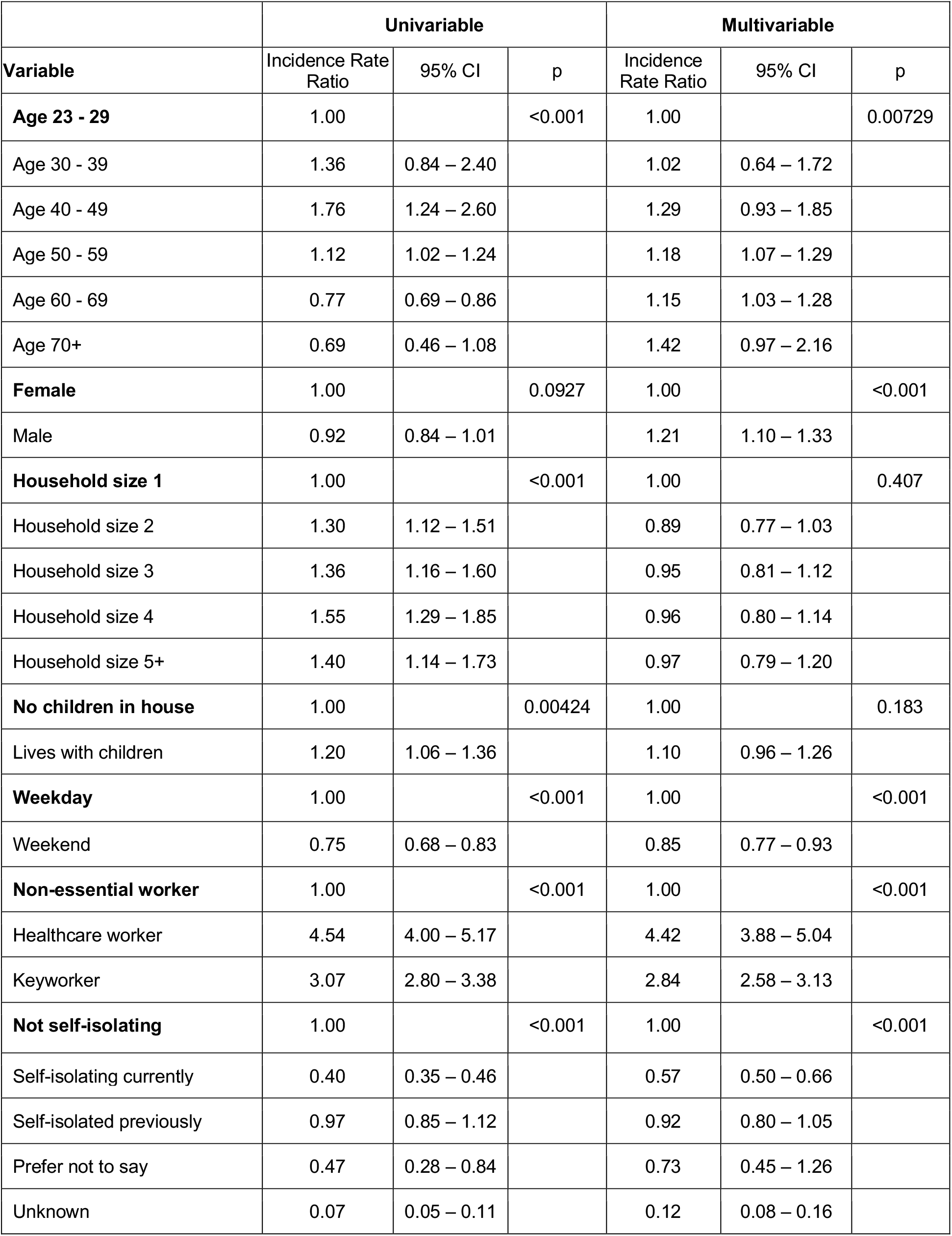
Relative number of daily total non-household contacts from univariable and multivariable negative binomial regression.

|  | Univariable |  |  | Multivariable |  |  |
| --- | --- | --- | --- | --- | --- | --- |
| Variable | Incidence Rate Ratio | 95% CI | p | Incidence Rate Ratio | 95% CI | p |
| **Age 23 - 29** | 1.00 |  | <0.001 | 1.00 |  | 0.00729 |
| Age 30 - 39 | 1.36 | 0.84 – 2.40 |  | 1.02 | 0.64 – 1.72 |  |
| Age 40 - 49 | 1.76 | 1.24 – 2.60 |  | 1.29 | 0.93 – 1.85 |  |
| Age 50 - 59 | 1.12 | 1.02 – 1.24 |  | 1.18 | 1.07 – 1.29 |  |
| Age 60 - 69 | 0.77 | 0.69 – 0.86 |  | 1.15 | 1.03 – 1.28 |  |
| Age 70+ | 0.69 | 0.46 – 1.08 |  | 1.42 | 0.97 – 2.16 |  |
| **Female** | 1.00 |  | 0.0927 | 1.00 |  | <0.001 |
| Male | 0.92 | 0.84 – 1.01 |  | 1.21 | 1.10 – 1.33 |  |
| **Household size 1** | 1.00 |  | <0.001 | 1.00 |  | 0.407 |
| Household size 2 | 1.30 | 1.12 – 1.51 |  | 0.89 | 0.77 – 1.03 |  |
| Household size 3 | 1.36 | 1.16 – 1.60 |  | 0.95 | 0.81 – 1.12 |  |
| Household size 4 | 1.55 | 1.29 – 1.85 |  | 0.96 | 0.80 – 1.14 |  |
| Household size 5+ | 1.40 | 1.14 – 1.73 |  | 0.97 | 0.79 – 1.20 |  |
| **No children in house** | 1.00 |  | 0.00424 | 1.00 |  | 0.183 |
| Lives with children | 1.20 | 1.06 – 1.36 |  | 1.10 | 0.96 – 1.26 |  |
| **Weekday** | 1.00 |  | <0.001 | 1.00 |  | <0.001 |
| Weekend | 0.75 | 0.68 – 0.83 |  | 0.85 | 0.77 – 0.93 |  |
| **Non-essential worker** | 1.00 |  | <0.001 | 1.00 |  | <0.001 |
| Healthcare worker | 4.54 | 4.00 – 5.17 |  | 4.42 | 3.88 – 5.04 |  |
| Keyworker | 3.07 | 2.80 – 3.38 |  | 2.84 | 2.58 – 3.13 |  |
| **Not self-isolating** | 1.00 |  | <0.001 | 1.00 |  | <0.001 |
| Self-isolating currently | 0.40 | 0.35 – 0.46 |  | 0.57 | 0.50 – 0.66 |  |
| Self-isolated previously | 0.97 | 0.85 – 1.12 |  | 0.92 | 0.80 – 1.05 |  |
| Prefer not to say | 0.47 | 0.28 – 0.84 |  | 0.73 | 0.45 – 1.26 |  |
| Unknown | 0.07 | 0.05 – 0.11 |  | 0.12 | 0.08 – 0.16 |  |

## DISCUSSION

In this paper, we demonstrate that social contacts dropped dramatically during the first lockdown in the UK, as measured in the ALSPAC cohort in 2020. Contact numbers were found to be much lower than pre-pandemic estimates, but exhibited a lower, non-zero, limit, associated with occupation, household size and presence of children living at home. This limit and its drivers partly explain patterns of transmission and COVID-19 reproduction rates during lockdown in the UK [22] [7]].

Social contact surveys have mainly been conducted as stand-alone studies by recruiting new participants [6] [7] [5]. Here, we surveyed an existing longitudinal cohort. This approach has several strengths. With ethics approval, consent and communications mechanisms in place it was possible to get a rapid assessment of mixing patterns. Participant engagement is high, evidenced by the high response rate. Using an existing cohort provides a more in-depth understanding of contacts and behaviours. Linkage with existing data provides the opportunity to understand social contact patterns in the context of other correlates such as occupation, household size, internet use, pet ownership, as well as subsequent follow-up to look at the long-term impact of COVID-19. Another example is the impact of COVID-19 on mental health, this has been explored in two longitudinal cohorts (ALSPAC and Generation Scotland) [23]. Young adults in the ALSPAC cohort were shown to experience a rise in magnitude of anxiety and reduction in well-being. The authors suggest this is possibly reflective of mitigation measures (i.e., lockdown and physical distancing) as opposed to risk of COVID-19 infection, with the latter determined as factor for higher depression and anxiety among older adults. Consequently, COVID-19 has differentially affected mental health in young and old adults, which may further be explained by contact patterns resulting from mitigation measures.

A limitation of the longitudinal cohort approach is that our survey is not demographically or geographically representative. As a birth cohort, participants were either born in 1991/1992 or they are parents of children born in those years, therefore we had few participants aged between 30 to 50 years old. Furthermore, the majority of participants are from the Bristol/Avon area and are less ethnically diverse than the UK as a whole. Given only 2% of the sample was non-white, and a further 9.4% were of unknown ethnicity, we were unable to investigate the association between ethnicity and contact patterns. However, evidence now strongly suggests that people from black, Asian, and minority ethnic (BAME) communities are disproportionately affected by COVID-19, with both racism and social determinants of health (e.g., high risk occupation, socioeconomic status, increased burden of comorbidities) identified as causes [24].

We acknowledge that the presented analyses are susceptible to collider bias since they have been restricted to ALSPAC participants who volunteered to respond to the COVID-19 questionnaire [25]. Participation in ALSPAC questionnaires by cohort participants has previously been demonstrated to be non-random; respondents are likely to be highly educated and health conscious (non-smokers) [26]. In addition, the COVID-19 questionnaire was completed online which may favour responses from those with internet access and engagement in technology [27] [28]. Consequently, the associations relating to the number of contacts could be biased due to colliders on participation. Ideally one would perform these associations using inverse probability weighting in which we estimate the probability of being selected into the sample using at least the exposure and outcome variables; however, this is not possible in our data because we do not have information on numbers of contacts outside of the selected sample. Instead, we reported the representativeness of the sample by comparing variable distributions outside the sample, and we observed that questionnaire participants were more likely to be female, white and aged 23–29 and 50–69. Similarly, analysis of variable distributions within the sample demonstrated that females, particularly younger females, and individuals with higher educational level (≥ A level) were more likely to respond [16]. Despite these differences in sample representation, the distribution of the number of contacts was comparable to pre-pandemic social contact surveys (right skewed, with a small fraction having the most contacts) [5] [29] [30].

Although we took an alternative approach to recruitment, our results are comparable to other social contact surveys. Compared to pre-pandemic surveys, we measured a significantly reduced number of social contacts per person. Our estimates are broadly consistent with other pandemic era surveys, such as CoMix, which reported an average of 2.8 contacts per person *(cf*. 3.7) at the height of the UK lockdown in May 2020 [5]. We modelled the distribution of contacts with a negative binomial model; a pre-pandemic contact survey found that a negative binomial distribution was unable to capture large groups [5], which are largely absent with physical distancing measures.

We observed striking occupational trends, with high numbers of contacts for healthcare workers and other keyworkers. Contacts of healthcare workers were more likely to involve touch, increasing their risk of exposure. The majority of keyworkers were identified as teachers in nursery, primary and secondary school settings, during a time of partial school closures. Interestingly, the group also included police officers. In line with contact surveys pre- and post-pandemic, essential contacts were higher for individuals living in larger households and with children [31] [6] [7]). Since these variables are related, associated contacts could be explained by lockdown placing pressures on parents and carers to seek non-household support, which necessarily involves contacts [32]. Finally, self-isolation was demonstrated to reduce daily contacts by 59%, suggesting that although effective in reducing contacts, some level of essential contacts outside of the household are maintained. These essential contacts may reflect financial pressures to continue work and/or caring responsibilities, necessity to obtain supplies or perceived risk of COVID-19 [32].

Understanding the drivers of contact patterns during the COVID-19 pandemic is informative for designing testing and vaccination strategies. For the latter, this will depend largely on whether a vaccine will interrupt transmission, prevent disease or both. As COVID-19 vaccines are rolled out in the UK and worldwide, priority groups are being determined; however, it is essential we look for evidence in informing these campaigns.

## Data Availability

Full details of the cohort and study design have been described previously and are available at http://www.alspac.bris.ac.uk.

Please note that the study website contains details of all the data that is available through a fully searchable data dictionary and variable search tool (http://www.bristol.ac.uk/alspac/researchers/our-data/).

ALSPAC data access is managed through a system of open access. The steps below highlight how to apply for access to the data included in this project (accessed under the project number B3514) and all other ALSPAC data:

1. Please read the ALSPAC access policy, which describes the process of accessing the data and samples in detail, and outlines the costs associated with doing so.
2. You may also find it useful to browse our fully searchable research proposals database, which lists all research projects that have been approved since April 2011.
3. Please submit your research proposal for consideration by the ALSPAC Executive Committee. You will receive a response within 10 working days to advise you whether your proposal has been approved.

Please note that a standard COVID-19 dataset will be made available at no charge (see description below); however, costs for required paperwork and any bespoke datasets required additional variables will apply.

The code used to conduct data analyses is available at https://github.com/amythomas/AlspacContactsQ1.git

## ACKNOWLEDGEMENTS

We are extremely grateful to all the families who took part in this study, the midwives for their help in recruiting them, and the whole ALSPAC team, which includes interviewers, computer and laboratory technicians, clerical workers, research scientists, volunteers, managers, receptionists and nurses. We are grateful to colleagues in the University of Bristol COVID-19 Emergency Research Group (UNCOVER) and ALSPAC COVID-19 Analysis Group for discussions supporting the manuscript. The UK Medical Research Council and Wellcome (Grant ref: 217065/Z/19/Z) and the University of Bristol provide core support for ALSPAC. This publication is the work of the authors and ACT, LD, HC, KN, DS, EN, AT, GH, SST, AF, NT, EBP will serve as guarantors for the contents of this paper. A comprehensive list of grants funding is available on the ALSPAC website (http://www.bristol.ac.uk/alspac/external/documents/grant-acknowledgements.pdf); This research was specifically funded by the Wellcome Trust, Medical Research Council and University of Bristol Elizabeth Blackwell Institute for Research (102215/2/13/2). ACT and GH are funded by the Wellcome Trust (217509/Z/19/Z and 208806/Z/17/Z). HC and EBP would like to acknowledge support from the National Institute for Health Research (NIHR) Health Protection Research Unit (HPRU) in Behavioural Science and Evaluation at the University of Bristol. HC is additionally funded through an NIHR Career Development Fellowship [CDF-2018-11-ST2-015]. The views expressed are those of the authors and not necessarily those of the NIHR or the Department of Health and Social Care. EN, LD and EBP are supported by the MRC (MR/V038613/1). LD and EBP are also supported by the MRC (MC/PC/19067) and LD is further supported by EPSRC (EP/V051555/1) and The Alan Turing Institute under the EPSRC grant EP/N510129/1. NJT is a Wellcome Trust Investigator (202802/Z/16/Z), is the PI of the Avon Longitudinal Study of Parents and Children (MRC & WT 217065/Z/19/Z), is supported by the University of Bristol NIHR Biomedical Research Centre (BRC-1215-2001), the MRC Integrative Epidemiology Unit (MC_UU_00011) and works within the CRUK Integrative Cancer Epidemiology Programme (C18281/A19169).

## DATA AVAILABILITY

Full details of the cohort and study design have been described previously and are available at http://www.alspac.bris.ac.uk. Please note that the study website contains details of all the data that is available through a fully searchable data dictionary and variable search tool (http://www.bristol.ac.uk/alspac/researchers/our-data/). ALSPAC data access is managed through a system of open access. The steps below highlight how to apply for access to the data included in this project (accessed under the project number B3514) and all other ALSPAC data:

1. Please read the <u>ALSPAC access policy</u>, which describes the process of accessing the data and samples in detail, and outlines the costs associated with doing so.
2. You may also find it useful to browse our fully searchable <u>research proposals database</u>, which lists all research projects that have been approved since April 2011.
3. Please <u>submit your research proposal</u> for consideration by the ALSPAC Executive Committee. You will receive a response within 10 working days to advise you whether your proposal has been approved.

## AUTHORS CONTRIBUTIONS

ACT, EBP and LD wrote the first draft of the manuscript. EBP, LD, NT and ACT had the original idea for the manuscript. ACT, LD, HC, KN, DS, EN, AT, GH, SST, AF, NT and EBP designed analyses, interpreted the results, and critically reviewed the manuscript. ACT, EBP and LD conducted the analyses.

