## Supplementary information for "Limits of lockdown: characterising essential contacts during strict physical distancing"

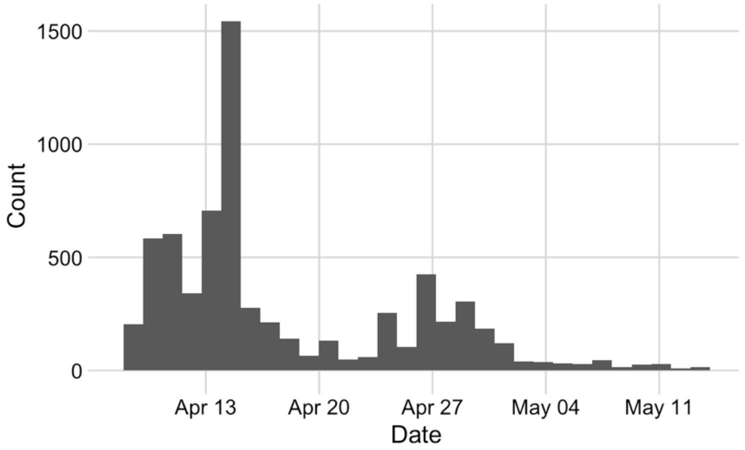


**Supplementary Figure 1: Frequency of questionnaire responses.** Responses received during the rollout period of 9th April 2020 to14th May 2020; peaks coincide with invitation and reminder emails

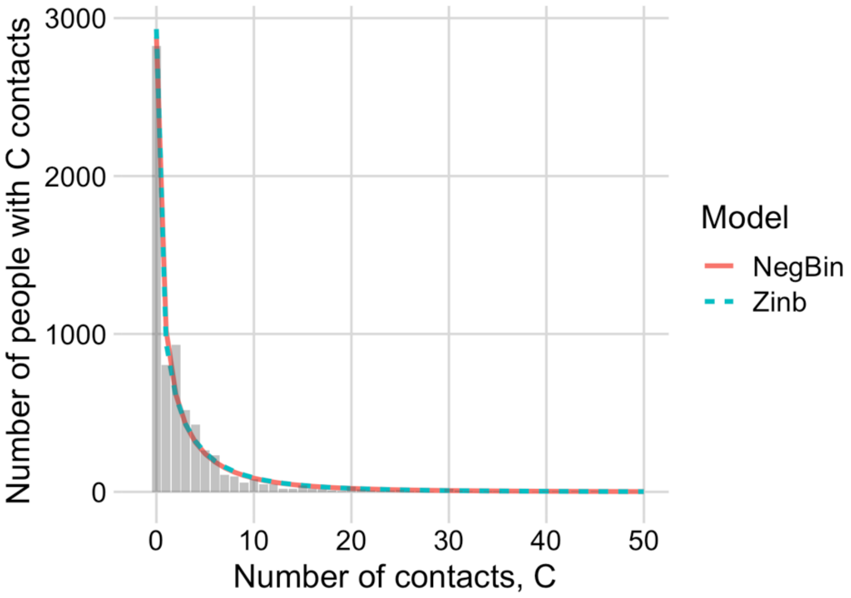


**Supplementary Figure 2: Model fit comparison.** Model fit plotted for negative binomial (NegBin), its zero inflated equivalent (Zinb) and observed counts. Proportion of observed zeros: 2825/6807 (41.5%); proportion of predicted zeros for negative binomial: 2881/6807 (42.3%) and zero inflated negative binomial: 2931/6807 (43.1%). Number of contacts truncated at 50.

**Supplementary Table 1: Characteristics of questionnaire participants in comparison to the South West and UK population.** Population estimates provided by ONS 2019 mid-year and 2011 census estimates

| | Questionnaire participants | | ONS South West estimate | ONS UK estimate |
|---|---|---|---|---|
| Characteristic* | n | (%) | (%) | (%) |
| Sex | 6807 | | | |
| Female | 4895 | 71.9 | 50.8 | 50.6 |
| Male | 1912 | 28.1 | 49.2 | 49.4 |
| Age† | 6807 | | | |
| 23–29 | 3039 | 44.6 | 12.9 | 13.1 |
| 30–39 | 45 | 0.7 | 16.1 | 18.8 |
| 40–49 | 90 | 1.3 | 16.4 | 17.8 |
| 50–59 | 2107 | 31.0 | 19.3 | 19.2 |
| 60–69 | 1455 | 21.4 | 16.4 | 15.0 |
| 70+ | 71 | 1.0 | 18.9 | 16.1 |
| Ethnicity‡ | 6177 | | | |
| Non-white | 137 | 2.2 | 4.7 | 12.9 |
| White | 6040 | 97.8 | 95.3 | 87.1 |
| Unknown | 630 | | | |

*Data for sex and age from the ONS 2019 mid-year estimates[1]

†No participants were aged under 23 years of age, percentages of age groups in the observed sample only were compared to ONS data. The 70+ age group was censored at 85.

‡Ethnicity data from the 2011 census for South West and UK populations[2]

### References

1. Office-for-National-Statistics. Office for National Statistics. Population estimates for the UK, England and Wales, Scotland and Northern Ireland: mid-2019, using April 2020 local authority district codes. 2021

2. Office-for-national-statistics. Census 2011. KS201 UK, Ethnic group. 2011.
